# Cohort-specific serological recognition of SARS-CoV-2 variant RBD antigens

**DOI:** 10.1101/2022.01.10.21268250

**Authors:** Douglas D. Fraser, Michael R. Miller, Claudio M. Martin, Marat Slessarev, Paul Hahn, Ian Higgins, Christopher Melo, Michael A. Pest, Nate Rothery, Xiaoqin Wang, Johannes Zeidler, Jorge A. Cruz-Aguado

## Abstract

**Background:** Estimating the response of different cohorts (e.g. vaccinated or critically ill) to new SARS-CoV-2 variants is important to customize measures of control. Thus, our goal was to evaluate binding of antibodies from sera of infected and vaccinated people to different antigens expressed by SARS-CoV-2 variants.

**Methods:** We compared sera from vaccinated donors with sera from four patient/donor cohorts: critically ill patients admitted to an intensive care unit (split in sera collected between 2 and 7 days after admission and more than ten days later), a NIBSC/WHO reference panel of SARS-CoV-2 positive individuals, and ambulatory or hospitalized (but not critically ill) positive donors. Samples were tested with an anti-SARS-CoV-2 IgG serological assay designed with microplates coated with a SARS-CoV-2 RBD recombinant antigen. The same sample sets were also tested with microplates coated with antigens harbouring RBD mutations present in eleven of the most widespread variants.

**Results:** Sera from vaccinated individuals exhibited higher antibody binding (P<0.001) than sera from infected (but not critically ill) individuals when tested against the WT and each of 11 variants’ RBD.

The optical density generated by sera from non-critically ill convalescence individuals upon binding to variant’s antigens was different (P<0.05) from that of the WT in some variants—noteworthy, Beta, Gamma, Delta, and Delta Plus variants.

**Conclusions:** Understanding differences in binding and neutralizing antibody titers against WT vs variant RBD antigens from different donor cohorts can help design variant-specific immunoassays and complement other diagnostic and clinical data to evaluate the epidemiology of new variants.

## Introduction

The race to understand the impact of new variants of the SARS-CoV-2 virus on the effectiveness of antibody therapeutics, vaccines, and infection-elicited antibody responses has led to an unprecedented number of converging studies, the majority showing that most potent ‘immune escape’ mutations are in the receptor-binding motif (RBM), a region located within the receptor binding domain (RBD) of SARS-CoV-2’s spike protein [1]. Indeed, approximately 90% of plasma or serum neutralizing antibody activity targets the RBD [2] and structural data support that RBD-based vaccines have a competitive position to deliver a fast response to the COVID-19 pandemic [3].

Until now, a trend has been delineated for the most frequent Variants of Concern (VOC) B.1.1.7 (Alpha), B.1.351 (Beta), P.1 (Gamma) and B1.617.2 (Delta) [4]. Some reports concur that sera from vaccinated or COVID-19 convalescent patients can efficiently neutralize viruses with just the N501Y mutation in the RBD, however, variants including the E484K mutation render sera from both wild type (WT) infected and vaccinated patients less efficient at virus neutralization [4-9]. Variants with sentinel mutation N439K decrease the activity of both polyclonal convalescent sera and monoclonal antibodies from individuals recovering from infection [10]. Another RBD mutation, L452R, present in variants B.1.429 and B.1.427 (Epsilon), and Indian Delta variant B.1.617.2 (Table 1), is thought to increase viral infectivity and potentially promote viral replication [11-15].

**Table 1.**
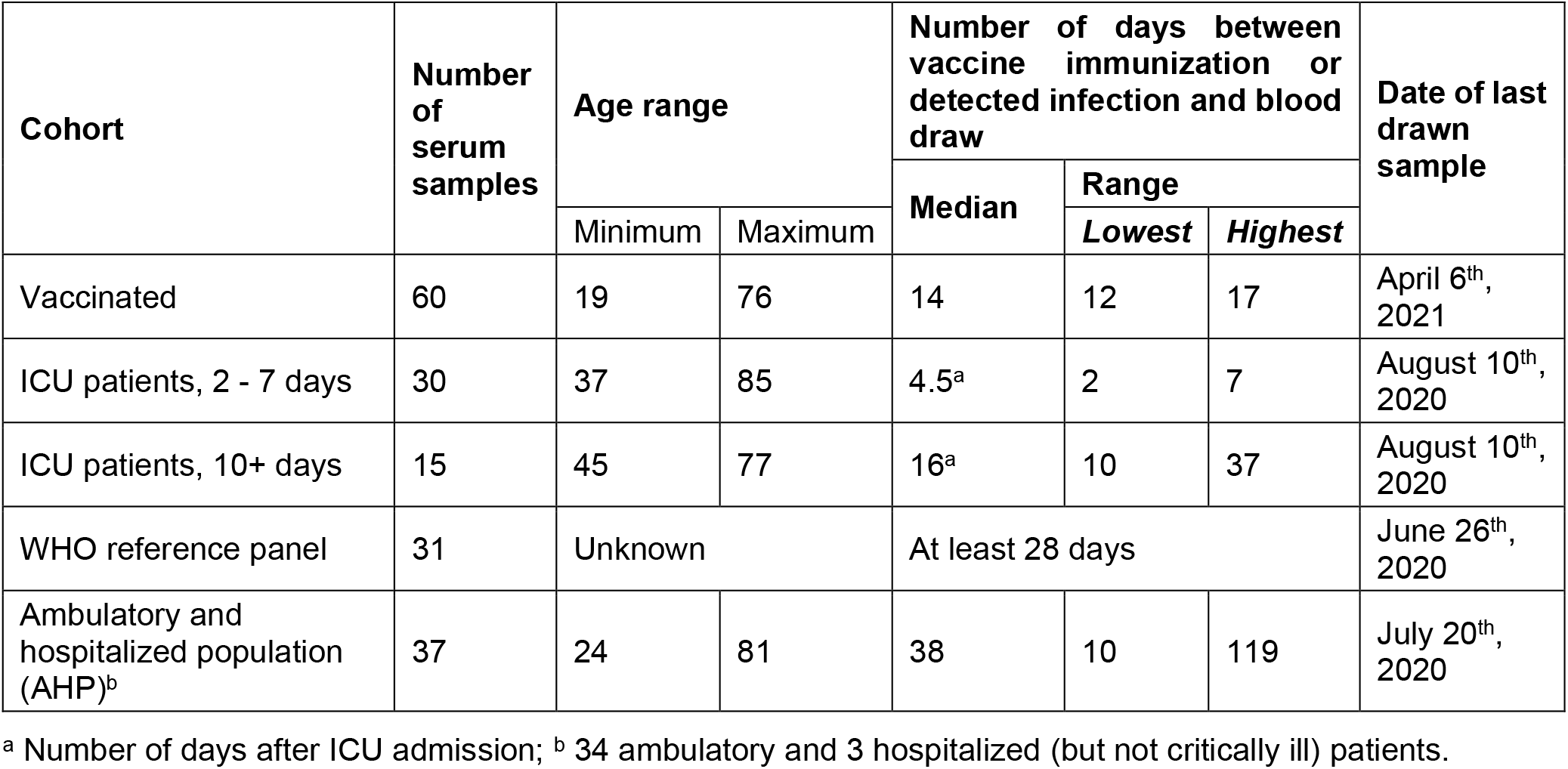
Summary of serum specimens used in this study. Additional patient data provided in Supplemental Material (Tables S1-S5).

The published research, however, has scarcely addressed differences between naturally infected individuals of the general population, critically ill hospitalized patients, and vaccinated subjects in the response to new variants.

Personalizing the response to new variants of the virus is important to optimize measures of control; for example, as precautionary warnings for travel within regions with a prevalent variant, or to better triage the selection of individuals who should be prioritized for vaccination. Although neutralization assays could provide answers to those questions, they are expensive and time consuming for practical testing of the population.

In this study, we set out to investigate first, how the recognition pattern of WT SARS-CoV-2 RBD antibodies generated by vaccination distinguishes those patients compared to the recognition pattern of individuals after infection, and furthermore, determine how this recognition pattern differs when these antibodies are presented with the mutated variant-associated SARS-CoV-2 RBD proteins. Secondly, we aimed to examine whether recognition of variant specific RBD differs between vaccine sera and antibodies produced by different post-infection cohorts, information that could complement clinical data with variant-specific immunoassay-based screenings to evaluate the potential epidemiological impact of new variants on previously infected and vaccinated cohorts.

## Methods

### Study population and Ethics

We used SARS-CoV-2 serum positive samples from:

a. **Donors vaccinated with mRNA vaccines**. Commercial samples from individuals who were vaccinated with two doses of either the Moderna mRNA-1273 or the Pfizer BioNTech COVID-19 vaccines during the first 3 months of 2021. The donor’s serum was collected before and after the first and second doses.
b. **Critically-ill patients** (split in sera collected between 2 and 7 days after admission and more than ten days later). Samples came from patients admitted to the level-3 academic ICU at London Health Sciences Centre (London, Ontario), confirmed as COVID-19 with standard hospital testing by detection of two SARS-CoV-2 viral genes using polymerase chain reaction [16].
c. **NIBSC/WHO reference panel of SARS-CoV-2 positive donors** from the NIBSC/WHO reference panels.
d. **Non-critically ill positive patient samples**. Commercial samples were collected during 2020. Donors were ambulatory or hospitalized patients (AHP) COVID-19 positive based on RT-PCR tests and immunoassays and tested again with the ELISA kit of this study. Three donors were hospitalized (not critically ill) and released.

Commercial samples were sourced through Access Biologicals (Vista, California, USA), Lampire Biological Laboratories (Pipersville, PA, USA), or Plasma Services Group, Inc (Moorestown, NJ, USA), each of which confirmed patient consent and participation in an Institutional Review Board (IRB) approved protocol. For critically ill patient recruitment, waived consent was approved for a short, defined period (Western University, Research Ethics Board [REB] number 1670). Samples obtained through the WHO database were originally collected under WHO protocols and ethical considerations, and specifics are publicly available online via links provided in S4. Ultimately, all patient samples were approved for research use by their respective sources. Furthermore, all samples were assigned arbitrary Sample ID’s to further anonymize personal data. Additional information available on request.

Samples were collected in North America before the global spread of the variants. We inferred that most of the studied positive specimens harbored antibodies raised against the WT virus, likely WA1/2020 (summarized in Table 1). More detailed information about each sample and the demographics, infection/vaccination timeline, and patient outcomes can be viewed in Supplemental Tables S1-S4.

### Immunoassays

Anti-SARS-CoV-2 IgG antibodies were detected with an enzyme-linked immunosorbent assay (ELISA) kit (DBC anti-SARS-CoV-2 ELISA, DBC-IGG-19) with interim authorization by Health Canada. In brief, the ELISA based assays were designed against antigens formulated by recombinant proteins (aa 319-541) created from the WT RBD sequence (original manufacturer’s ELISA design) or those representing the RBD of prevalent VOCs (Table 2) with their various mutation sets (N501Y, K417N-E484K-N501Y, K417T-E484K-N501Y, L452R, L452R-E484Q, L452R-T478K, K417NL452R-T478K, N439K, Y453F, S477N, K417T).

**Table 2.** Classification of the variants and RBD mutations associated with the antigens of this study.

| Variant name | Lineage (PANGO) | Clade (GISAID) | RBD mutations | Country of first detection |
| --- | --- | --- | --- | --- |
| Wild Type | B | G | - | China |
| Alpha | B.1.1.7 | GRY | N501Y | United Kingdom |
| Beta | B.1.351/501Y.V2 | GH | K417N, E484K and N501Y | South Africa |
| Gamma | P.1/B.1.1.28.1. | GR | K417T, E484K, and N501Y | Brazil |
| Epsilon | B.1.427/429 | GH | L452R | Denmark |
| Kappa | B.1.617.1 | G | L452R, E484Q | India |
| Delta | B.1.617.2 | G | L452R, T478K | India |
| Delta Plus | B.1.617.3 | G | K417N, L452R, T478K | India |
| N439K | B.1.141/B.1.258 | GR | N439K | United Kingdom |
| Y453F | N/A | N/A | Y453F | Denmark |
| S477N | B.1.526.2/B1.1.25 | 20.C | S477N | Australia |
| K417T | N/A | N/A | K417T | Brazil |

The original WT assay (CAN-IGG-19) uses a ratio between the optical density (OD) of the sample and the cut-off **[Ratio = OD of sample / Cut-Off]** to determine a ‘positive’ vs ‘negative’ result, where the Cut-Off **[Cut-Off (CO) = (Mean of 3 Negative Control results) x factor 1.5]** is used to generate a ratio which is then interpreted as a Positive (Ratio ≥ 1.2), Negative (Ratio ≤ 1.0), or Borderline (Ratio 1.0 – 1.2) result.

In this study, the ratio was used only to compare the results between cohorts within the same antigen. When comparisons were made between antigens, we used the optical density (OD) generated by the binding of samples’ antibodies to the antigens (rather than the ratio); in this way isolating the antigen-antibody interaction and avoiding bias generated by differences in binding between the antigens and the negative control.

More details on the layout and performance of the original test can be found in [16] and in the kit’s IFU (https://dbc-labs.com/products/elisa/anti-sars-cov-2-igg/).

DiaSorin Liaison SARS-CoV-2 S1/S2 IgG results were provided by the sample supplier.

### Neutralization antibodies study

To establish the validity of the DBC-IGG-19 ELISA kit to detect neutralizing antibodies we ran a comparison study against Genscript cPass™ SARS-CoV-2 Neutralization Antibody Detection/Surrogate Virus Neutralization Test Kit (NJ, USA). This assay was previously validated against plaque reduction neutralization tests PRNT50 and PRNT90 with 100% agreement (See manual for SARS-CoV-2 Surrogate Virus Neutralization Test Kit).

The samples were run with both the Genscript Kit and the DBC ELISAs for each of the WT and variants listed above following the instructions for use of each test.

### Variant-specific immunoassays

To test the binding of antibodies to mutated antigens, microplates were coated with recombinant RBD harbouring mutations present in 11 of the most widespread variants of the virus to date (Table 2). The expression system used to generate the mutant antigen RBD was identical to that of the previously released DBC kit (DBC-IGG-19) with the only difference being the mutations themselves. The mutated antigens were coated under the same conditions used for the WT antigen currently in the commercial anti-SARS-CoV-2 IgG immunoassay. All other reagents of the variant kits used and serological test conditions remained the same as the WT unaltered assay (DBC-IGG-19).

### Statistical Analysis

Statistical analysis was performed using Analyse-it software (Analyse-it Software, Ltd. (UK); https://analyse-it.com/). Univariate group comparisons with non-parametric Steel pairwise ranking were calculated to compare variants within each cohort (figure 2) and cohorts within each variant (figure 3). P-values at the 5% significance level were used to establish differences between variables.

We compared the results of WT antigen to antigens with RBD mutations within each patient cohort based on optical density (OD) which represents binding between antibodies in patient samples and the presented antigen (i.e. the antigen coated onto the microplates), rather than ratios because the ratios are calculated using the negative control as a reference of the cut-off. Therefore, the ratios are affected by changes in the binding of the Negative control to the mutated antigens (Figure 2). To compare cohorts within each variant we used the ratio, because as long as the same antigen is used, normalizing by the negative control enables assessment of the clinical result (positive, borderline, or negative) (Figure 3).

## Results

### Detection of neutralizing antibodies

To evaluate the capacity of the RBD variant DBC ELISA kits in this study to detect neutralizing antibodies we compared patient sample results against the Genscript cPass™ SARS-CoV-2 Neutralization Antibody Kit.

All 168 samples that generated a positive neutralizing antibody result with the GenScript test were also confirmed with the DBC ELISA for 100% positive agreement (See supplemental Table S5, borderline results are counted as positive). We observed a positive exponential relationship between the inhibition rate produced by the GenScript kit (x) and the ratio generated by the ELISA (y), y = 0.4275e0.0385x, r = 0.85 (Figure 1).

**Fig 1.**
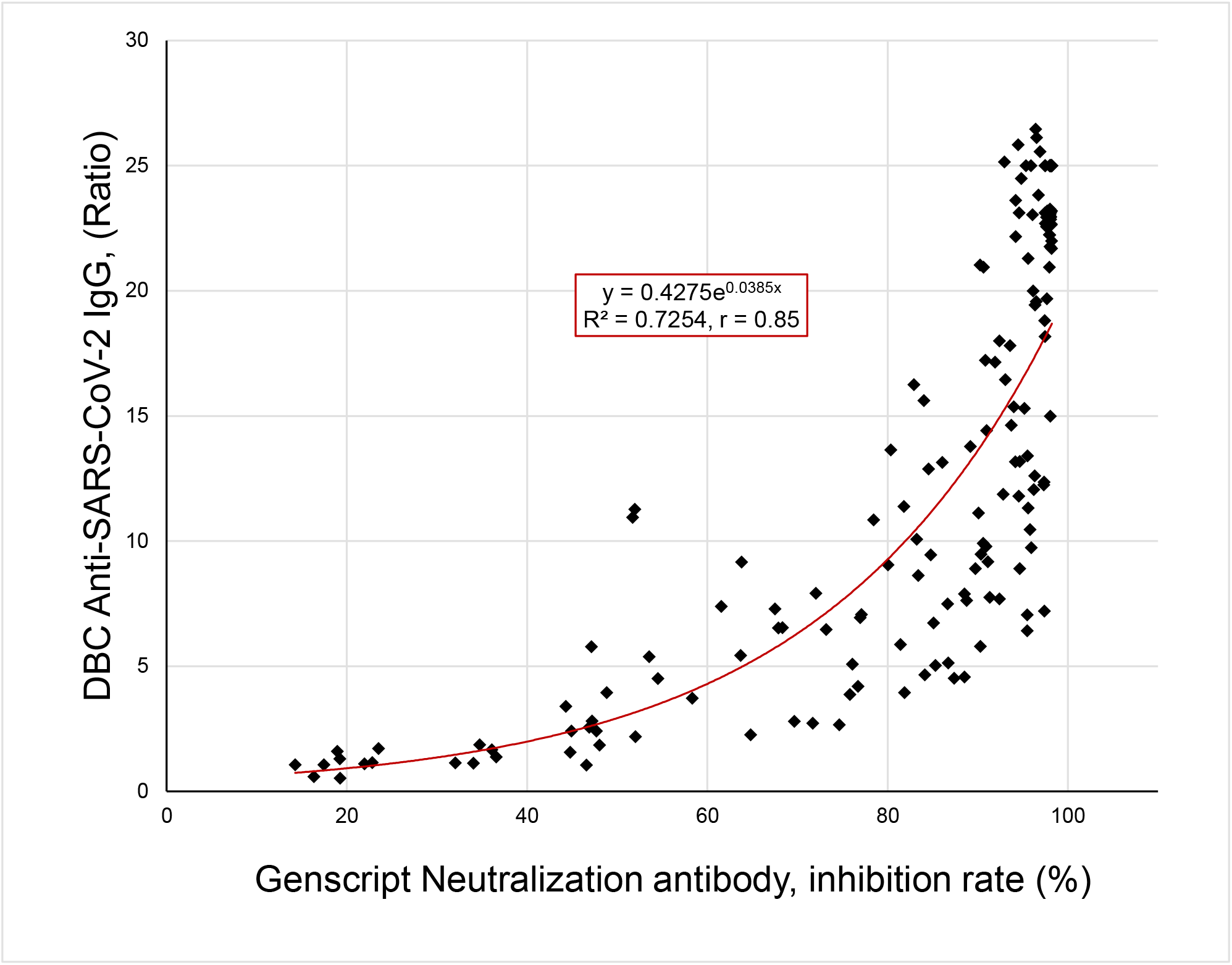
Relationship between neutralization and IgG antibody ratio as determined by the Genscript cPass™ SARS-CoV-2 Neutralization Antibody Detection/Surrogate Virus Neutralization Test Kit (L00847) and DBC’s Health Canada authorized anti-SARS-CoV-2 IgG (CAN-IGG-19). Ratio (DBC kit) was calculated based on a Cut Off [Cut-Off (CO) = (Mean of 3 Negative Controls) x factor 1.5] which is then used to generate a ratio [Ratio = OD of sample / Cut-Off].

Additionally, the GenScript neutralizing antibody kit reported a positive result in 98.2% (165 of 168) of COVID seroconverters also detected by the DBC ELISA (counting borderline DBS results as negative). The few seropositive samples that produced a negative neutralizing antibody result had a low serological positive ratio (< 1.9) relatively close to the Cut-off.

### Reactivity of vaccine and infection sera with WT and mutated antigens

To assess how the humoral immune response in vaccinated individuals compares to antibodies generated from WT infection, and the further influence of the mutations present in the RBD of SARS-CoV-2 variants, we purchased commercial samples of sera from individuals who had received two doses of the Moderna mRNA-1273 or the Pfizer BioNTech COVID-19 vaccines during the first 3 months of 2021.

Even though the donors recruited for the study had declared themselves healthy and uninfected with COVID-19, 25% had a positive pre-vaccine anti-SARS-CoV-2 IgG result with the DBC assay (18% with DiaSorin, Tables S1 and S2 in Supplemental Material). In 80% of those donors, the ELISA signal was above the maximal OD of 4 after the first vaccine dose suggesting that the asymptomatic infection might have been equivalent to a first vaccine dose.

For the rest of the vaccinated donors, the ELISA ratio increased more than two times between the first and the second vaccination doses. After the second vaccination dose, binding responses appear to converge, as we no longer observed a higher SARS-CoV-2 IgG ELISA ratio among the subjects who had a viral infection before the vaccination. In fact, only four (including one sample from a donor positive before the vaccination) of 60 samples did not produce a ratio higher than 15 after two vaccination doses (Supplemental Table S1). We, therefore, estimate that a prior infection had a negligible effect in the load of anti-SARS-CoV-2 antibodies after two vaccine doses

Sera from vaccinated individuals were compared to sera of three COVID-19 convalescent cohorts: a) ICU patients, split into short (2-7 days) and longer (10+ days) hospitalization, b) a reference panel of samples from SARS-CoV-2 positive donors produced by NIBSC/WHO (WHO), and c) infected ambulatory or hospitalized population (AHP) samples.

For AHP and WHO samples, the ELISA OD response of the serological tests in four of the variants was not different (p>0.05) from WT antigen (Figure 2)—all four variants had a single mutation in the RBD (N501Y, L452R, K417T, or S477N). This pattern was different for the vaccinated, for whom three of the above single mutations and two triple mutation antigens displayed no differences in relation to the WT antigen (Figure 2). No significant differences (p>0.05) were observed between the WT and variants in the reactivity of ICU sera (Figure 2). We observed a consistent trend across individual donors (Supplemental Figure S1).

**Figure 2.**
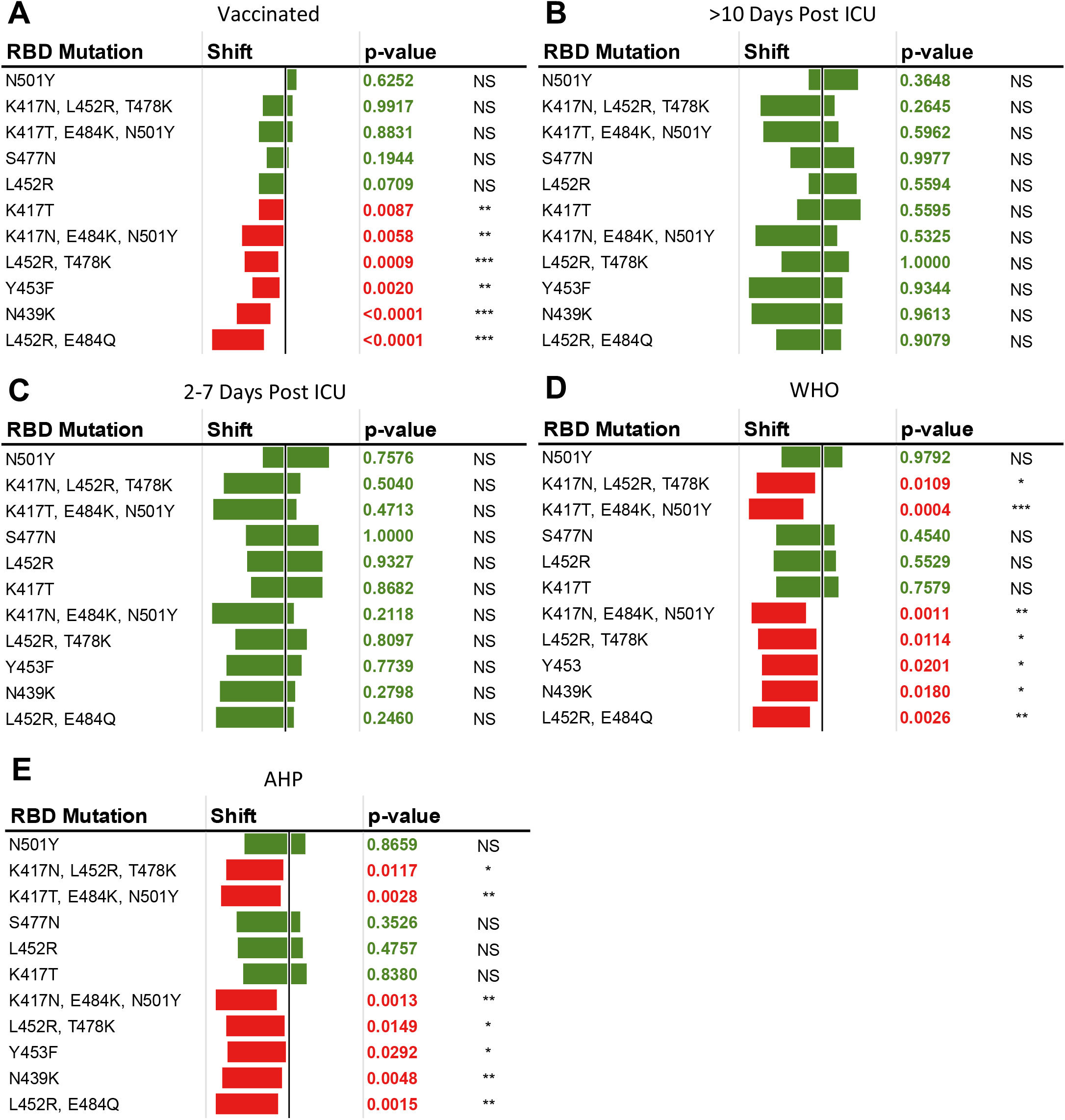
Comparative evaluation of antibody recognition of mutated antigens against WT in five cohorts by optical density (OD). The OD, rather than the ratio, was used to prevent bias from differences in the ELISAs’ negative control OD between antigens. Univariate Steel pairwise ranking non-parametric comparisons against the WT antigen. Experimental datasets for cohorts (A) Vaccinated, (B) >10 days post ICU admission, (C) 2-7 days post ICU admission, (D) WHO (E) Ambulatory or hospitalized population (AHP), (NS) nonsignificant, (^*^) P<0.05; (^**^) P<0.01, (^***^) P<0.001.

Sera from vaccinated individuals consistently exhibited a higher ratio (p<0.001) than sera from infected (but not critically ill) individuals (AHP, WHO) when tested against the WT and each of the 11 variants (Figure 3). Only the vaccinated cohort displayed a median ratio higher than 15 against all the mutated antigens—except the N439K antigen, which significantly lowered the median of ratio results in all cohorts. Still, even against this antigen, none of the samples from vaccinated donors produced a negative ratio result (<1.0). (Figure 3).

**Figure 3.**
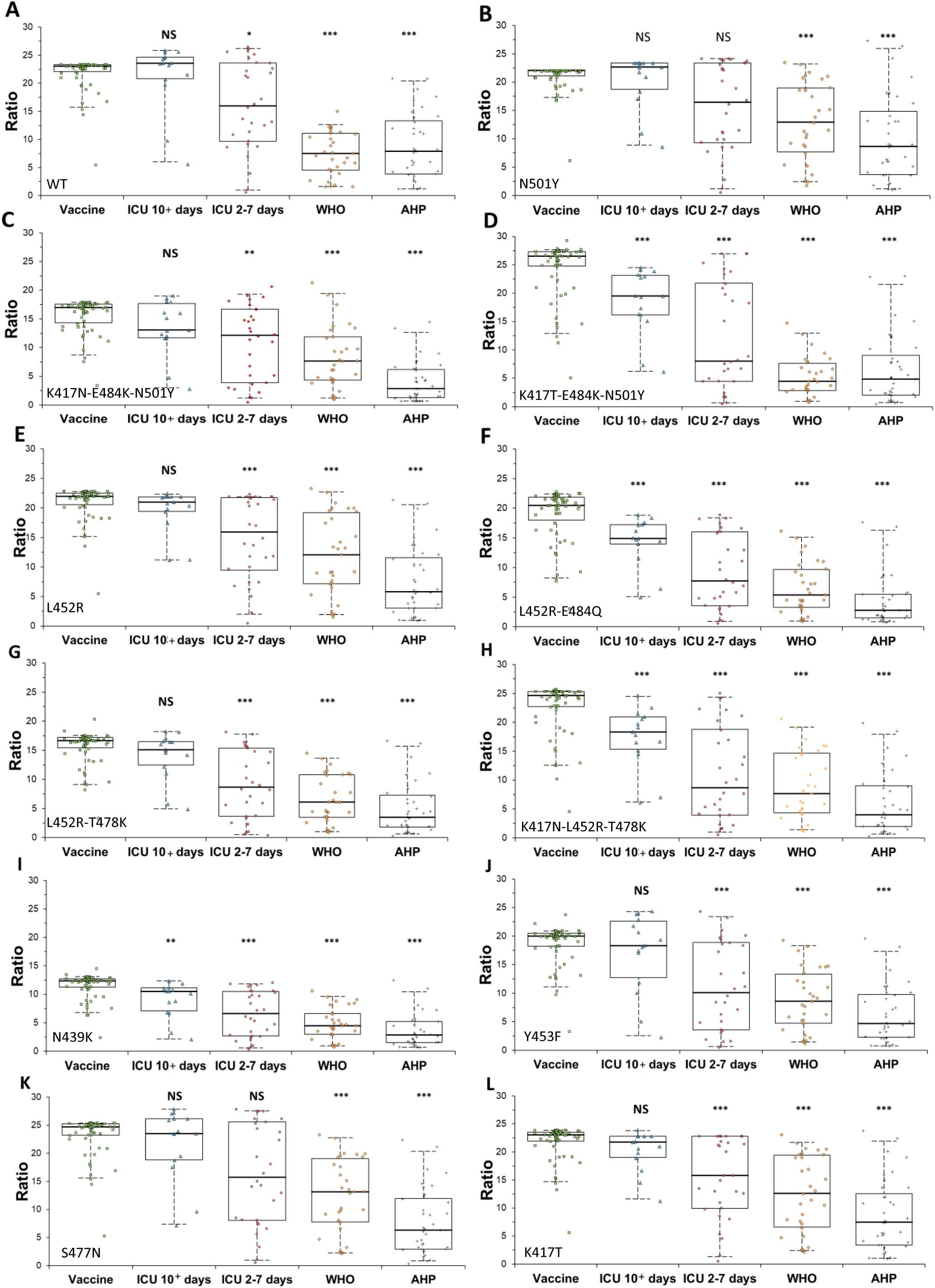
Reactivity of sera from vaccinated individuals (Vaccine), from patients 2-7 (ICU 2-7 days) and more than 10 days (ICU 10+ days) after admission to ICU, from a NIBSC/World Health Organization (WHO) panel, and from ambulatory and hospitalized population (AHP) to recombinant antigens comprising the RBD of wild type (WT) SARS-CoV-2 or recombinant antigens with mutations present in variants of the virus. Ratio was calculated based on a Cut Off [Cut-Off (CO) = (Mean of 3 Negative Controls) x factor 2.0] which is then used to generate a ratio [Ratio = OD of sample / Cut-Off]. The cohorts were compared against the vaccinated using Steel pairwise ranking non-parametric method: (NS) nonsignificant, (^*^) P<0.05; (^**^) P<0.01, (^***^) P<0.001. The median is plotted as a line. The box represents 1^st^ to 3^rd^ quartiles and the whiskers the minimum and maximum of the 95% confidence interval. **(A)** WT, **(B)** N501Y, **(C)** K417N-E484K-N501Y, **(D)** K417T-E484K-N501Y, **(E)** L452R, **(F)** L452R-E484Q, **(G)** L452R-T478K, **(H)** K417N-L452R-T478K, **(I)** N439K, **(J)** Y453F, **(K)** S477N, **(L)** K417T.

The second highest antibody response was from critically ill ICU patients who were hospitalized for more than 10 days. For seven of twelve of the antigens, including the WT antigen, this cohort was not different from the vaccinated. ICU patients upon admission (2-7 days), WHO, and AHP samples presented lower positive ratios (Figure 3).

To assess if the differences between the cohorts were due to lower age range of vaccinated and AHP in comparison to ICU patients (Table 1), we split the ratio results of the vaccinated and AHP cohorts into those from donors younger and older than 45 (since the ICU 10+ were all older than 45) and compared them. We found no age-related differences (p>>0.05) (Supplemental Figures S2, S3).

## Discussion

Differences in cross-variant seroreactivity between population cohorts, including vaccinated individuals and severely ill patients, have been scarcely documented. Current concerns are that some variants might escape neutralizing antibodies (an issue just recently developing with the recent multi-mutation bearing variant Omicron), but it is unclear how factors such as vaccines and the severity of the disease can influence the outcome. In this study, we used sera from vaccinated and previously infected individuals to evaluate their ability to recognize mutations in the SARS-CoV-2 RBD. We show that vaccine sera exhibited higher reactivity than convalescence sera from non-critically ill individuals against all twelve studied SARS-CoV-2 RBD antigens tested, including the WT RBD, while the reactivity of critically ill sera bridged those cohorts in most variants.

Understanding the nature of viral and vaccine-induced immunity is critical to managing the course of an epidemic [17]; in particular, how individuals infected early in the pandemic, or those who have been vaccinated will be protected against emerging variants, more importantly, variants holding mutations in the RBD—a region targeted by ∼90% of the neutralizing antibody activity [2].

To establish the capability of the ELISA used in this study to detect neutralizing antibodies we compared its performance against the Genscript cPass SARS-CoV-2 Neutralization antibody detection kit and found 100% positive agreement. This performance was likely enabled by the fact that the antigen in the ELISA comprises the critical RBD region of the virus. Indeed, immunoassays that use the full spike as antigen show a diminished ability to detect neutralizing antibodies [18].

Having established the capacity of the serological test to detect neutralizing antibodies, we proceeded to evaluate how sera from COVID infected critically ill patients recognize antigens harbouring RBD mutations. Previously, we have shown that IgG levels increase soon after admission to the ICU [16]. Here, we found that sera from ICU patients did not significantly distinguish any of the mutated antigens when compared to WT (p>0.05, Figure 2). Perhaps, severe, and prolonged infection induces a broad polyclonal antibody variability enabling reactivity to a wide range of antigenic variants.

For other cohorts, including the vaccinated, not all the mutations caused a decrease in the antigen’s antibody recognition (Figure 2). Vaccine sera did not discriminate five of the variants against the WT according to ELISA OD values, while WHO and AHP samples showed that four of the mutated target antigens, all bearing a single mutation, enabled the same level of antigen recognition in comparison to the WT. Conversely, mutations N439K (present in deemed extinct lineages B1.141 and B1.258), K417N-E484K-N501Y (Beta variant), and L452R-T478K (Delta variant) produced lower ODs (p<0.05) than the WT across the vaccinated, WHO and AHP panels. Differences between vaccine sera and WHO or AHP sera might be expected since the antigen configuration and display generated by the mRNA vaccines is dramatically different from that of the live viruses [3]. For example, K417T-E484K-N501Y (Beta variant) and L452R-E484Q (Delta Plus variant) were distinguished from the WT by WHO and AHP cohorts but not by the vaccinated.

It is noteworthy that the WHO and AHP panels, comprised of samples collected from two separate and unrelated populations, yielded almost identical results (Figure 2). Both panels identified the antigens with mutations corresponding to three VOCs (Beta, Gamma, Delta) Delta Plus, a fact that suggests that variant-specific immunoassay-based screenings on existing sera samples could serve as prospective tests to assess the potential impact of new variants before sufficient epidemiological data is available.

However, we believe that our results do not necessarily extrapolate to patterns in the *in vivo* neutralization of variants because we are examining antigens constructed solely by the RBD, while the mutations’ effect on virus affinity to the ACE2 receptor is not considered here. Nor are we examining the influence of variants’ full set of concomitant mutations outside of the RBD. Nevertheless, considering that the RBD plays a critical role in the antigenicity of new variants [2] we hypothesized that a relationship might exist between patient vulnerability to infection and disease and the reactivity of antibodies to isolated variant RBD. This is important considering that in a short time most of the worlds’ population should have some degree of immunity either by infection or vaccines’ (57% of the world’s population received at least one COVID-19 vaccine dose by December 23^rd^ 2021, according to Ourworldindata.org).

Our results tend to match previous findings. Compared to the WT, neutralization of B.1.1.7 (Alpha) and P.1 (Gamma) was found to be roughly equivalent [19], while neutralization of B.1.351 (Beta) spike protein was lower but still relatively robust. Neutralization titers against mutant viruses containing key spike mutations: N501Y and E484K-N501Y-D614G were found to be 1.46 and 0.81 in relation to the WT virus respectively [4] which matches well with the results we obtained for WHO and AHP panels for those mutations (Figure 2). Similarly, Wu et al. [5] detected reductions of the titer of neutralizing antibodies by a factor of 1.2 with pseudo-viruses encoding the B.1.1.7 (Alpha) variant compared to a factor of 6.4 and 3.5 against the B.1.351 (Beta) and P.1 (Gamma) variants. Those studies again align with our data showing that antibody binding to antigen with N501Y mutation alone and WT antigen are the same in all tested cohorts. Conversely, mutants including E484K-N501Y produced lower binding in WHO and AHP samples. Other reports, however, indicate that both B.1.1.7 (Alpha) and B.1.351 (Beta) lead to a decrease in neutralization [9,14,20].

Epidemiological data is conflicting but tend to assign higher mortality to mutations associated with B.1.351 (Beta) than to B.1.1.7 (Alpha) [21]. However, a recent study found increased transmissibility of variants that included mutations N501Y and E484K but not increased disease severity [22].

Residues N501 and E484 play different functions in the infectivity of the virus. Residue N501 is at the RBD-ACE2 interface and mutation N501Y was found to result in an increase of affinity to ACE2 [23]. Mutation E484, in turn, has been identified as an immunodominant spike protein residue with various mutations, including E484K, supporting escape from several monoclonal antibodies [24]. This divergence of functions between residues N501 and E484 might explain why we found E484K to reduce binding of WT-induced antibodies whereas N501Y and WT did not differ.

Additionally, in studies with monoclonal antibodies, the spike’s B.1.1.7 (Alpha) mutations were shown to reduce neutralization mostly of antibodies specific to the spike’s amino-terminal domain (NTD) but only with a small proportion of RBD specific antibodies [25].

Considering the immunodominance of the RBD, this could explain some of the moderate reduction in neutralizing activity of convalescence sera against authentic B.1.1.7 (Alpha) or pseudo-viruses carrying the B.1.1.7 (Alpha) spike mutations [26, 27] and the lack of diminished seroreactivity to N501Y by any of the cohorts of this study (Figure 2).

Furthermore, neutralization by some RBD-specific and NTD-specific monoclonal antibodies was found to be unaffected by variation in the spike protein suggesting the presence of cross-neutralizing epitopes in both the RBD and NTD [28].

Remarkably, mutation N439K produced one of the most consistent drops in ODs compared to WT and other variants (Fig 2). Early in 2021, N439K was the second most prevalent mutation of the RBD sequence [29], but currently is not one of the top ten most distributed mutations worldwide. This mutation is noteworthy because it enhances the binding affinity for the ACE2 receptor and decreases the neutralizing activity of both monoclonal antibodies and serum polyclonal antibodies of convalescence patients [10].

However, a deep mutational scanning (DMS) study did not find that mutation N439K significantly alters neutralization by polyclonal antibodies in plasma [30] in contrast to the findings described above [10] and our results (Figs 2 and 3). According to Harvey et al. [31], this discrepancy derives from the fact that the mechanism of immune escape provided by N439K is based on increased affinity to ACE2 rather than by directly affecting epitope recognition. Perhaps then, the experimental design of the DMS study is less sensitive to detecting immune evasion mutations of this type—an inconsistency that exposes limitations of DMS [32] and highlights the significance of testing the ability of immune (COVID-19 positive) sera to recognize new variants with real human samples and serological-based assays.

Mutation S477N—that has emerged several times during the pandemic [31]—was found in one study to be resistant to neutralization by a panel of monoclonal antibodies, but by contrast, responds similarly to the WT when tested with convalescence (polyclonal) serum [33]—a result that aligns with the data of this study (Figure 2), once again underscoring the advantage of testing real patient sera to evaluate the antigenicity of new mutations.

A recent study found lineage B.1.617.2 (Delta) to be associated with an increase in disease severity [34]. This variant also caused a higher rate of vaccine breakthrough cases (17.4% compared to 5.8% for all other variants) in Texas, with 8.4% of all COVID-19 cases occurring in fully vaccinated individuals [35]. In our study, the Delta and kappa variants (which share the L452R mutation) eluded vaccine and infection sera antibodies more than the WT in all cohorts except for critically ill patients (Figure 2).

However, in vaccinated individuals, we observed a shift towards higher ODs when mutation K417N was added to the Delta variant mutations L452R and T478K to replicate the Delta Plus variant (B1.617.3, K41N-L452R-T478K) mutation set. These additional mutations diminished the decreased binding seen in the Delta variant compared to Wild Type (p<0.001), which ultimately demonstrated a similar binding profile for the Delta Plus variant vs Wild Type (Figure 2). Not much epidemiology data have been collected so far about the Delta Plus variant, but it has been pointed out that this variant spreads more easily and is potentially more infectious [36].

As the recently emerging Omicron variant begins to spread, it is relevant to this study to note that this VOC bears 5 of the spike protein mutations examined (N501Y, K417N, E484A, T478K, S477N) [37]. While it is beyond the scope of this study to evaluate the effects of these mutations individually or collectively in the immunoreactivity of the Omicron variant, we believe evaluation of this VOC and future inevitable variants using similar assay panels may give insight into immune escape effects.

Our study has some limitations. We have not investigated how antibodies generated by vaccines other than the mRNA based Moderna and Pfizer products would respond to variant antigens; vaccines with different antigen presentations might result in a different pattern of variant recognition. The study is also limited by the fact that we were not able to examine how immune sera collected several months or years after infection or vaccination would affect recognition of RBD in antigens of new variants.

## Conclusions

In summary, our results indicate that:

a. Recognition of SARS-CoV-2 RBD in the sera of vaccinated individuals is significantly enhanced compared to sera from non-critically infected patients regardless of the antigen variant.
b. The antibodies generated in critically ill individuals are less variant-specific than those of non-critically ill and vaccinated subjects.
c. The antibodies present in the sera of non-critically ill convalescent donors distinguish some variants— noteworthy, Beta, Gamma, Delta, and Delta Plus variants—in relation to the WT, a fact that could enable variant-specific immunoassay-based screenings to aid evaluating the potential epidemiological impact of new variants.

## Data Availability

All data produced in the present study are available upon reasonable request to the authors.

## SUPPLEMENTAL MATERIAL

**Table S1.**
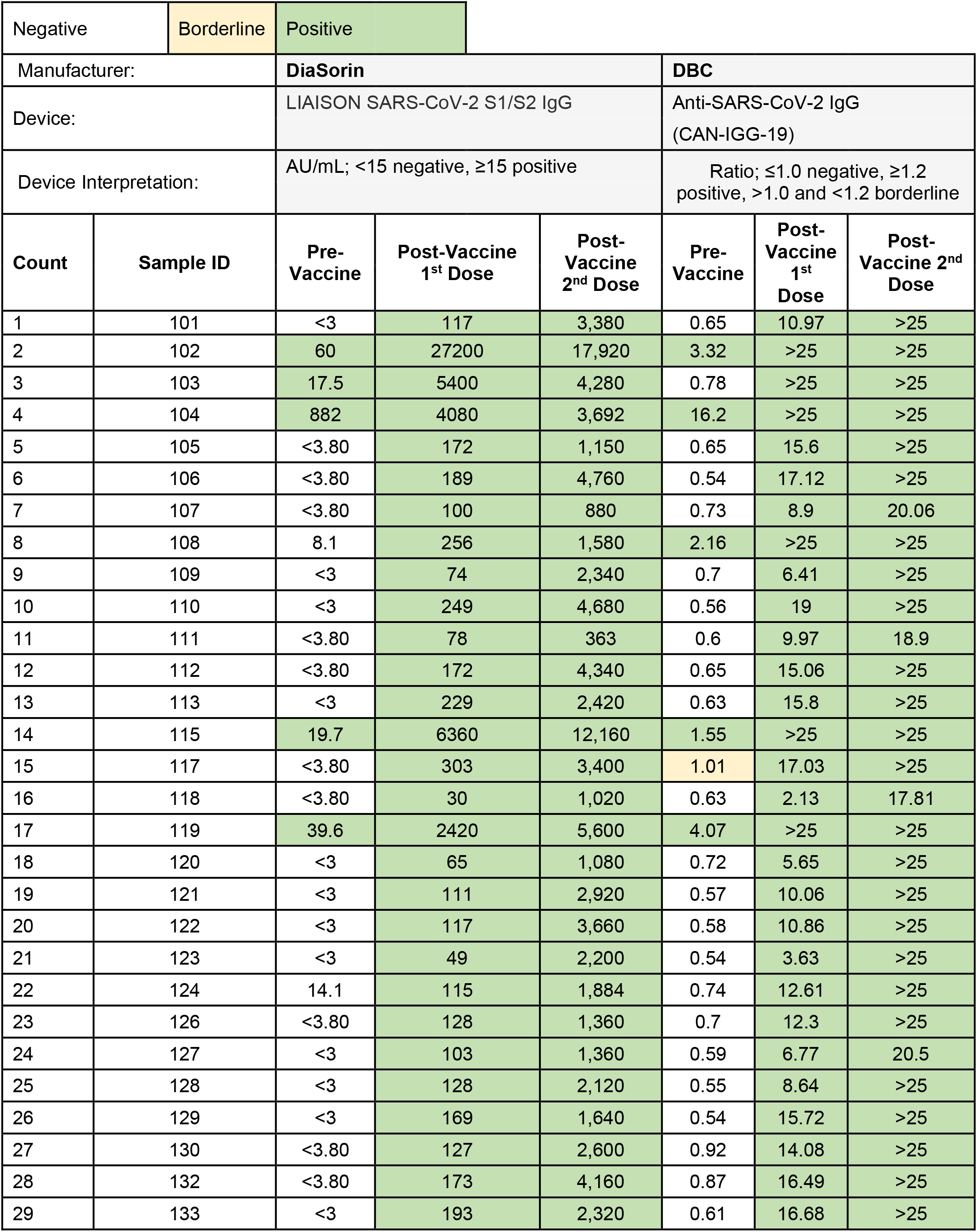
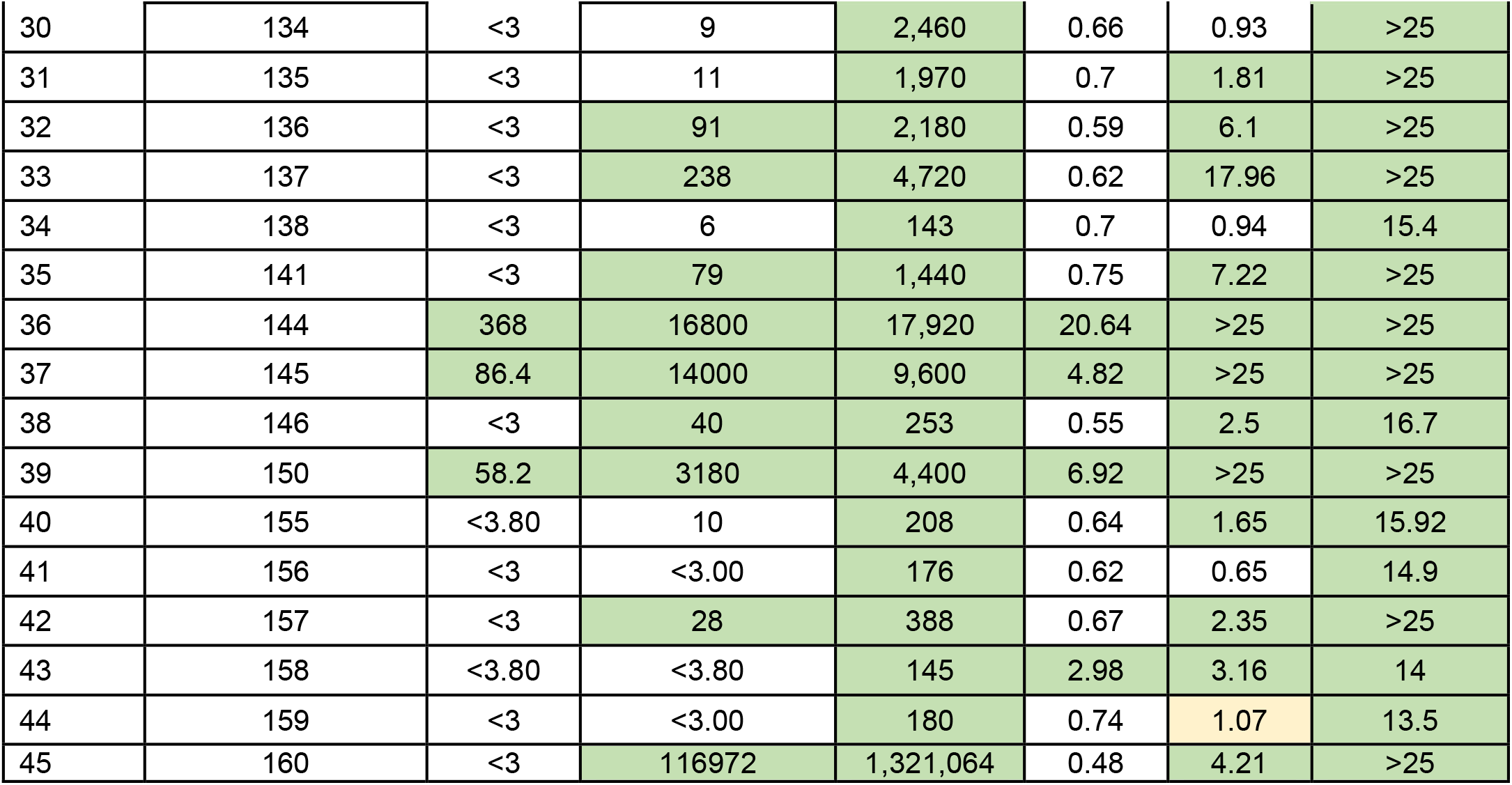
Analysis of samples from the Moderna COVID-19 Vaccine Serum Panel

**Table S2.** Analysis of samples from the Pfizer COVID-19 Vaccine Serum Panel.

| Negative |  | Borderline |  | Positive |  |  |  |
| --- | --- | --- | --- | --- | --- | --- | --- |
| Manufacturer: |  | DiaSorin |  |  | DBC |  |  |
| Device: |  | LIAISON SARS-CoV-2 S1/S2 IgG |  |  | Anti-SARS-CoV-2 IgG<br>(CAN-IGG-19) |  |  |
| Device Interpretation: |  | AU/mL; <15 negative, ≥15 positive |  |  | Ratio; ≤1.0 negative, ≥1.2 positive, >1.0 and <1.2 borderline |  |  |
| Count | Sample ID | Pre-Vaccine | Post-Vaccine 1st Dose | Post-Vaccine 2nd Dose | Pre-Vaccine | Post-Vaccine 1st Dose | Post-Vaccine 2nd Dose |
| 1 | 114 | 117 | 5,200 | 4,880 | 6.85 | >25 | >25 |
| 2 | 116 | 11.1 | 127 | 1,120 | 2.48 | 15.47 | 18.2 |
| 3 | 125 | 131 | 1,430 | 1,090 | 16.87 | >25 | 18.1 |
| 4 | 131 | <3.80 | 81 | 1,350 | 0.76 | 7.84 | >25 |
| 5 | 139 | <3.80 | 23 | 169 | 1.39 | 3.53 | 14.02 |
| 6 | 140 | <3.80 | 98 | 50 | 0.74 | 9.89 | 4.82 |
| 7 | 142 | <3.80 | 7 | 2,920 | 0.72 | 1.51 | >25 |
| 8 | 143 | <3.80 | 36 | 2,150 | 0.62 | 2.63 | >25 |
| 9 | 147 | <3.80 | 25 | 226 | 0.5 | 1.95 | 16.7 |
| 10 | 148 | <3.80 | 102 | 4,080 | 0.58 | 9.16 | >25 |
| 11 | 149 | <3.80 | 25 | 2,070 | 0.84 | 1.91 | >25 |
| 12 | 151 | <3.80 | 34 | 1,640 | 0.7 | 3.58 | >25 |
| 13 | 152 | 12.8 | 3,468 | 4,280 | 1.61 | >25 | >25 |
| 14 | 153 | <3.80 | 33 | 261 | 0.53 | 4.21 | 18.54 |
| 15 | 154 | 100 | 7,760 | 8,240 | 6.63 | >25 | >25 |

**Table S3.** Age and sex of sample donor in the 4 Cohorts

| **Vaccinated** |  |  |
| --- | --- | --- |
| **Sample #** | **Age** | **Sex** |
| 101 | <20 | M |
| 102 | 21-30 | F |
| 103 | 21-30 | F |
| 104 | 21-30 | F |
| 105 | 21-30 | M |
| 106 | 21-30 | M |
| 107 | 31-40 | F |
| 108 | 31-40 | F |
| 109 | 31-40 | F |
| 110 | 31-40 | F |
| 111 | 31-40 | F |
| 112 | 31-40 | F |
| 113 | 31-40 | F |
| 114 | 31-40 | F |
| 115 | 31-40 | M |
| 116 | 31-40 | M |
| 117 | 41-50 | F |
| 118 | 41-50 | F |
| 119 | 41-50 | F |
| 120 | 41-50 | F |
| 121 | 41-50 | F |
| 122 | 41-50 | F |
| 123 | 41-50 | F |
| 124 | 41-50 | F |
| 125 | 41-50 | F |
| 126 | 41-50 | M |
| 127 | 41-50 | M |
| 128 | 41-50 | M |
| 129 | 41-50 | M |
| 130 | 41-50 | M |
| 131 | 41-50 | M |
| 132 | 51-60 | F |
| 133 | 51-60 | F |
| 134 | 51-60 | F |
| 135 | 51-60 | F |
| 136 | 51-60 | F |
| 137 | 51-60 | F |
| 138 | 51-60 | F |
| 139 | 51-60 | F |
| 140 | 51-60 | F |
| 141 | 51-60 | M |
| 142 | 51-60 | M |
| 143 | 51-60 | M |
| 144 | 61-70 | F |
| 145 | 61-70 | F |
| 146 | 61-70 | F |
| 147 | 61-70 | F |
| 148 | 61-70 | F |
| 149 | 61-70 | F |
| 150 | 61-70 | M |
| 151 | 61-70 | M |
| 152 | 61-70 | M |
| 153 | 61-70 | M |
| 154 | 61-70 | M |
| 155 | 71-80 | F |
| 156 | 71-80 | F |
| 157 | 71-80 | F |
| 158 | 71-80 | M |
| 159 | 71-80 | M |
| 160 | 71-80 | M |
| **AVE** | **50.1** |  |
| **Median** | **49.5** |  |
| **SD** | **14.8** |  |
| **Females** |  | **38** |
| **Males** |  | **22** |

| **AHP** |  |  |
| --- | --- | --- |
| **Sample #** | **Age** | **Sex** |
| 161 | 21-30 | F |
| 162 | 21-30 | F |
| 163 | 21-30 | F |
| 164 | 21-30 | F |
| 165 | 21-30 | M |
| 166 | 31-40 | F |
| 167 | 31-40 | F |
| 168 | 31-40 | F |
| 169 | 31-40 | F |
| 170 | 31-40 | F |
| 171 | 31-40 | F |
| 172 | 31-40 | M |
| 173 | 31-40 | M |
| 174 | 41-50 | F |
| 175 | 41-50 | F |
| 176 | 41-50 | F |
| 177 | 41-50 | F |
| 178 | 41-50 | F |
| 179 | 41-50 | F |
| 180 | 41-50 | M |
| 181 | 41-50 | M |
| 182 | 51-60 | F |
| 183 | 51-60 | F |
| 184 | 51-60 | F |
| 185 | 51-60 | F |
| 186 | 51-60 | F |
| 187 | 51-60 | F |
| 188 | 51-60 | F |
| 189 | 51-60 | F |
| 190 | 51-60 | M |
| 191 | 51-60 | M |
| 192 | 51-60 | M |
| 193 | 51-60 | M |
| 194 | 61-70 | F |
| 195 | 61-70 | F |
| 196 | 71-80 | F |
| 197 | 81-90 | F |
| **AVE** | **46.9** |  |
| **Median** | **47** |  |
| **SD** | **13.5** |  |
| **Females** |  | **28** |
| **Males** |  | **9** |

| ICU 2 – 7 days |  |  |  |
| --- | --- | --- | --- |
| Sample # | Age | Sex | Outcome |
| 198 | 31-40 | M | ALIVE |
| 199 | 41-50 | F | ALIVE |
| 200 | 41-50 | M | ALIVE |
| 201 | 41-50 | M | ALIVE |
| 202 | 41-50 | M | DEAD |
| 203 | 41-50 | M | ALIVE |
| 204 | 51-60 | F | ALIVE |
| 205 | 51-60 | F | ALIVE |
| 206 | 51-60 | F | ALIVE |
| 207 | 51-60 | F | ALIVE |
| 208 | 51-60 | M | ALIVE |
| 209 | 51-60 | M | ALIVE |
| 210 | 51-60 | M | ALIVE |
| 211 | 51-60 | M | ALIVE |
| 212 | 51-60 | M | ALIVE |
| 213 | 61-70 | F | ALIVE |
| 214 | 61-70 | F | ALIVE |
| 215 | 61-70 | F | ALIVE |
| 216 | 61-70 | F | ALIVE |
| 217 | 61-70 | F | ALIVE |
| 218 | 61-70 | M | ALIVE |
| 219 | 61-70 | M | ALIVE |
| 220 | 61-70 | M | ALIVE |
| 221 | 61-70 | M | ALIVE |
| 222 | 71-80 | F | ALIVE |
| 223 | 71-80 | M | DEAD |
| 224 | 71-80 | M | ALIVE |
| 225 | 71-80 | M | ALIVE |
| 226 | 81-90 | F | DEAD |
| **AVE** | **60.7** |  |  |
| **Median** | **61** |  |  |
| **SD** | **11.3** |  |  |
| **Females** |  | **12** |  |
| **Males** |  | **17** |  |

| ICU 10+ days |  |  |  |
| --- | --- | --- | --- |
| Sample # | Age | Sex | Outcome |
| 227 | 41-50 | F | ALIVE |
| 228 | 41-50 | M | ALIVE |
| 229 | 41-50 | M | ALIVE |
| 230 | 51-60 | F | ALIVE |
| 231 | 51-60 | F | ALIVE |
| 232 | 51-60 | F | ALIVE |
| 233 | 51-60 | F | ALIVE |
| 234 | 51-60 | M | ALIVE |
| 235 | 61-70 | F | ALIVE |
| 236 | 61-70 | F | ALIVE |
| 237 | 61-70 | F | ALIVE |
| 238 | 61-70 | M | ALIVE |
| 239 | 61-70 | M | ALIVE |
| 240 | 71-80 | F | ALIVE |
| **AVE** | **58.3** |  |  |
| **Median** | **56** |  |  |
| **SD** | **8.5** |  |  |
| **Females** |  | **9** |  |
| **Males** |  | **6** |  |

**Table S4.**
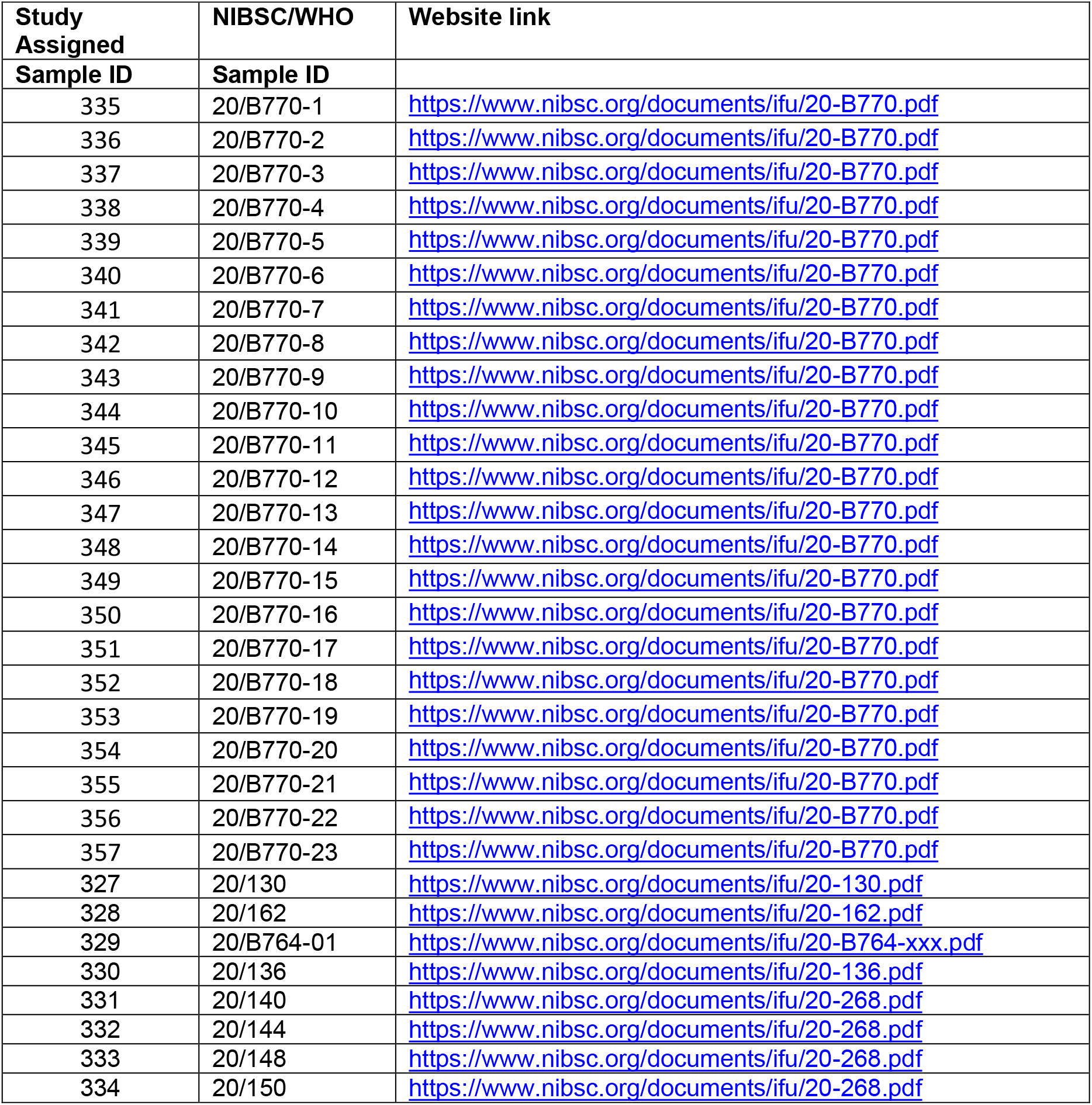
Links to sample NIBSC/WHO information.

**Table S5.**
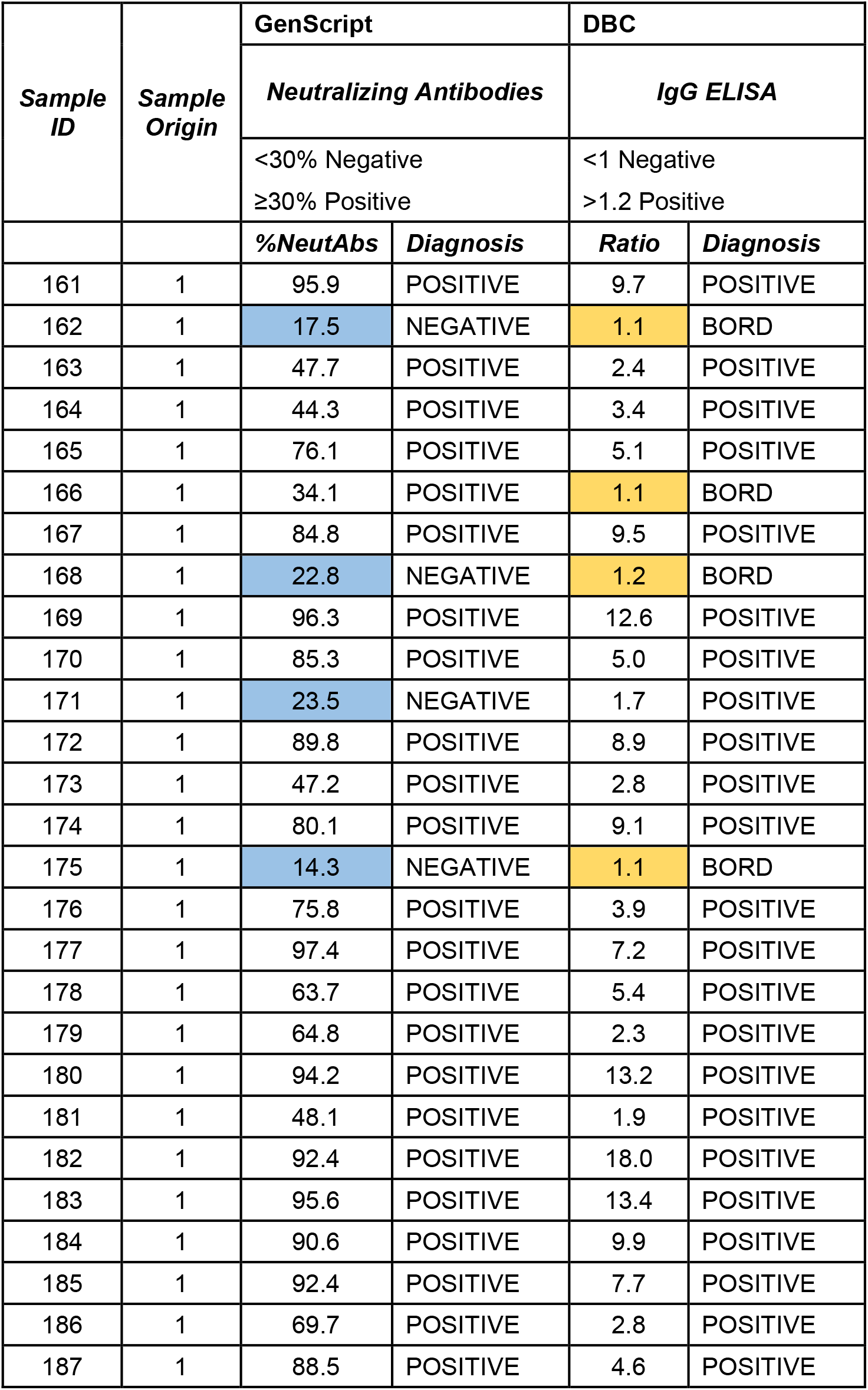
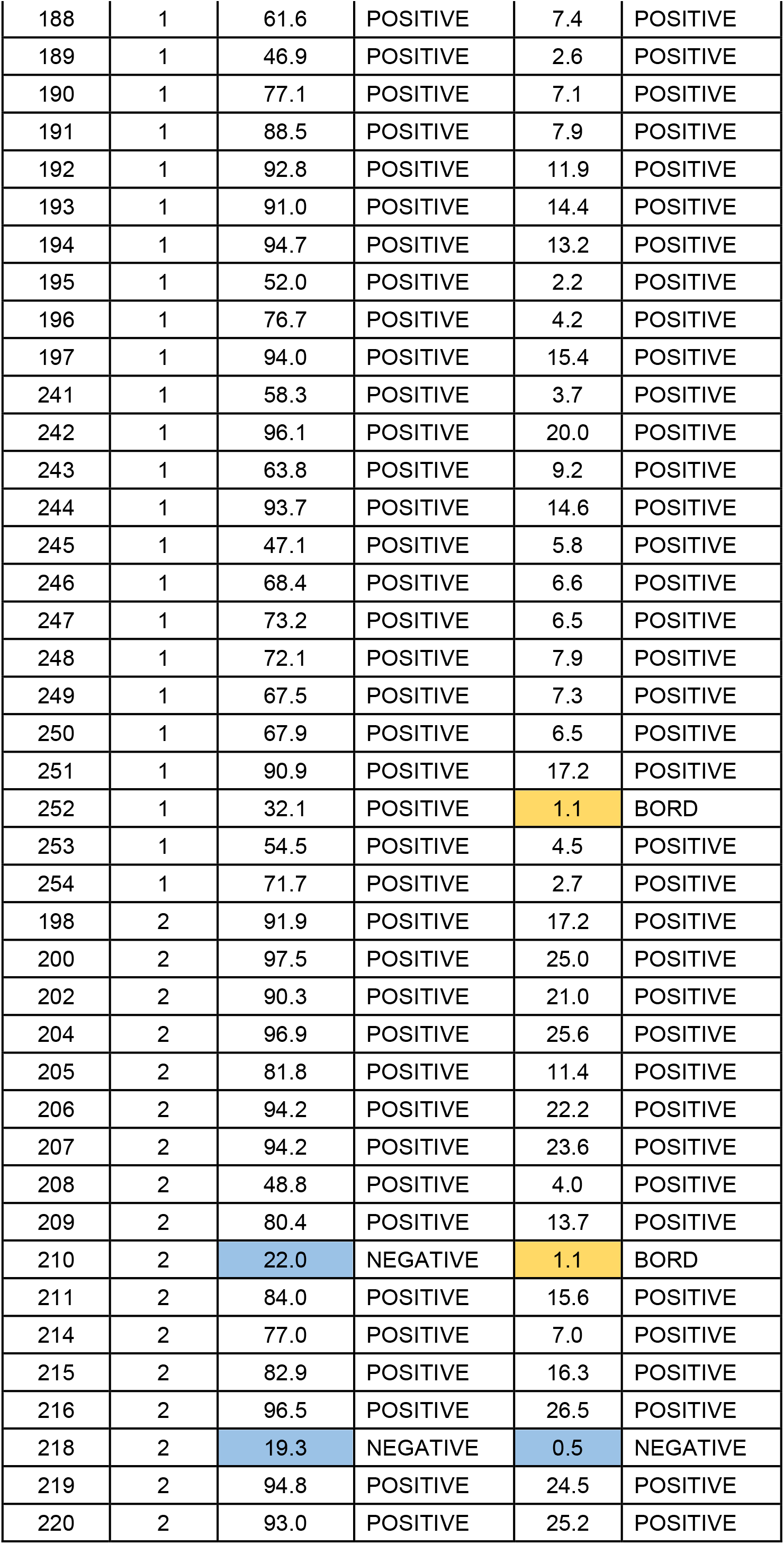
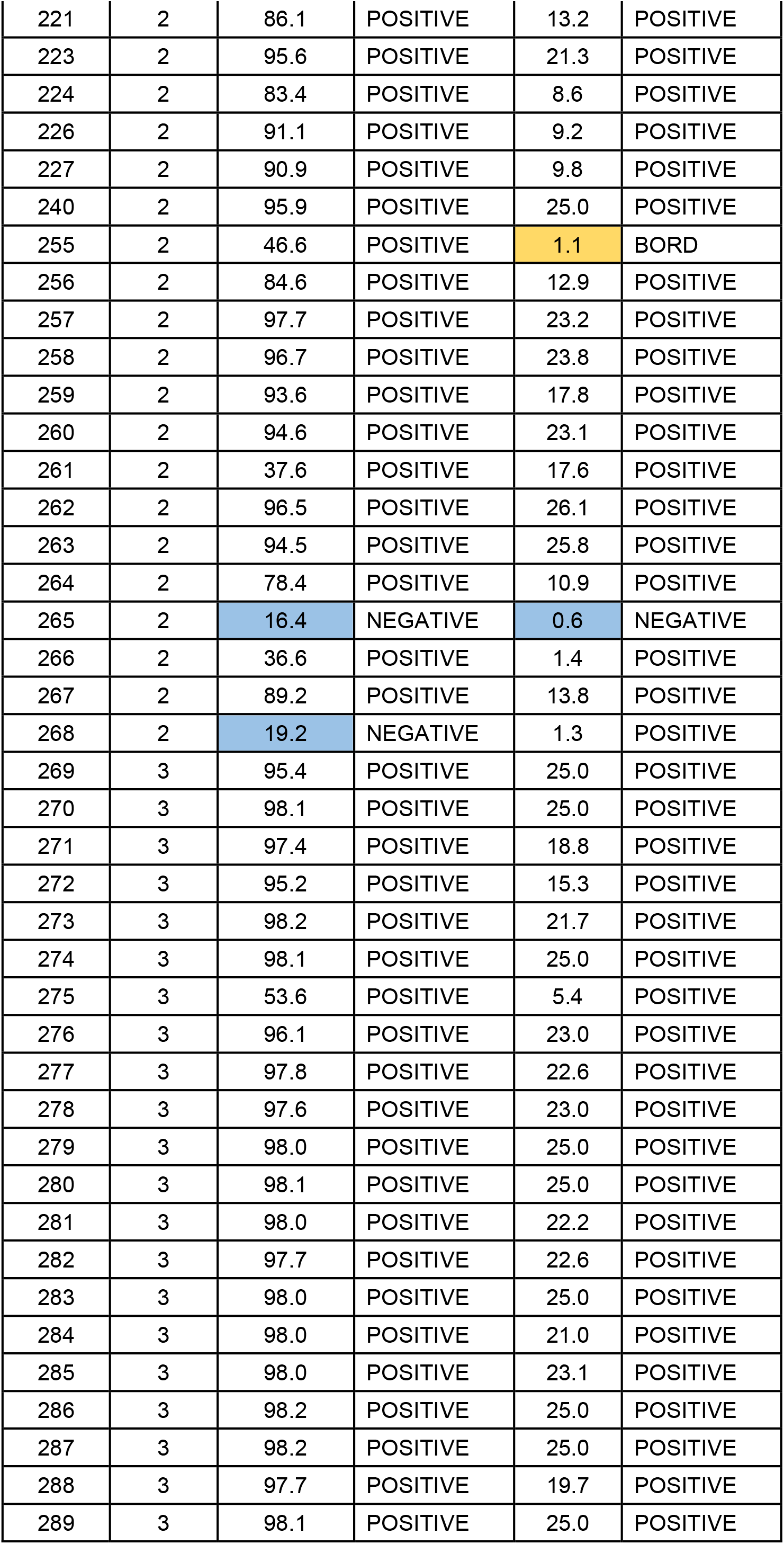
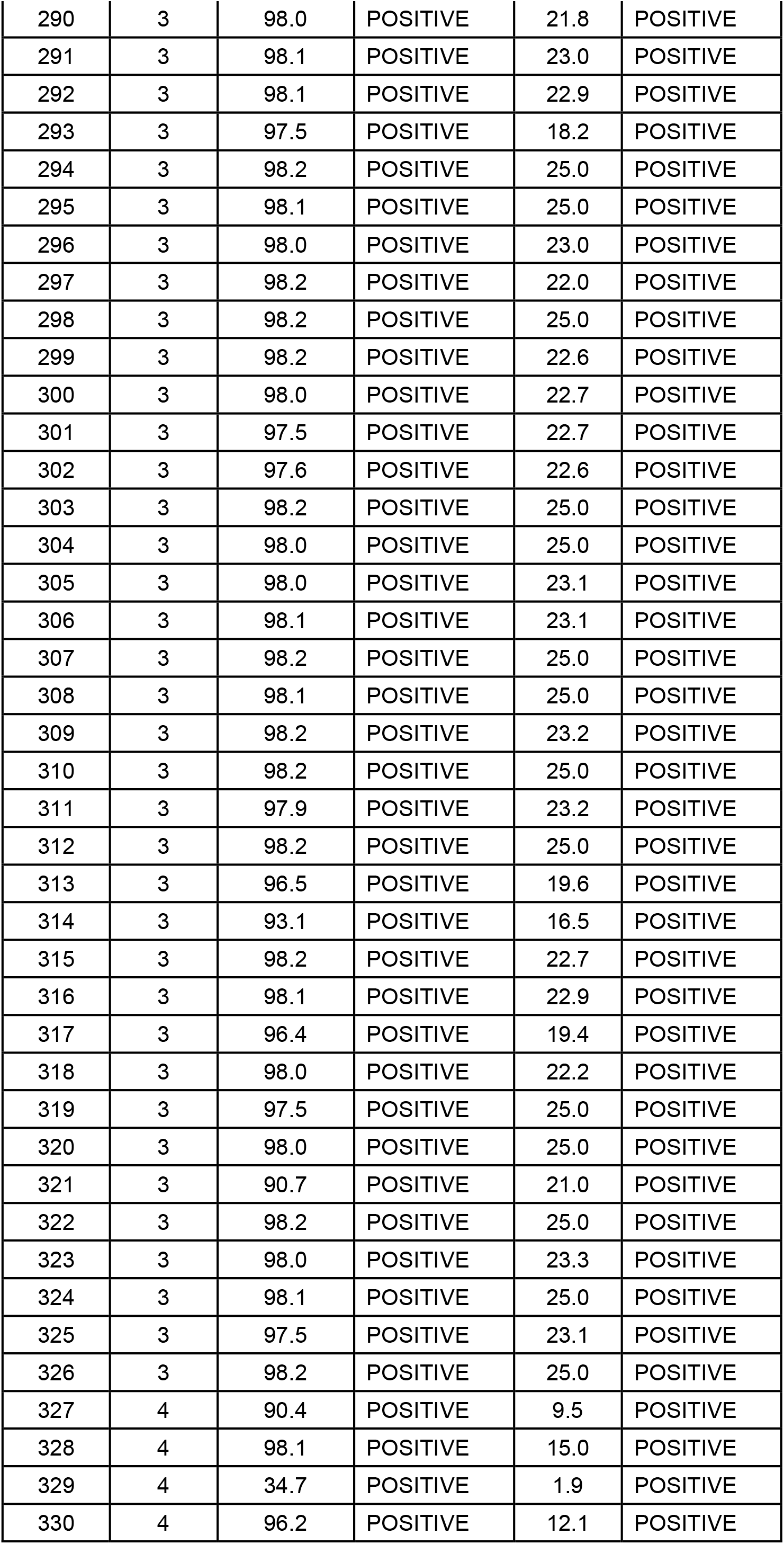
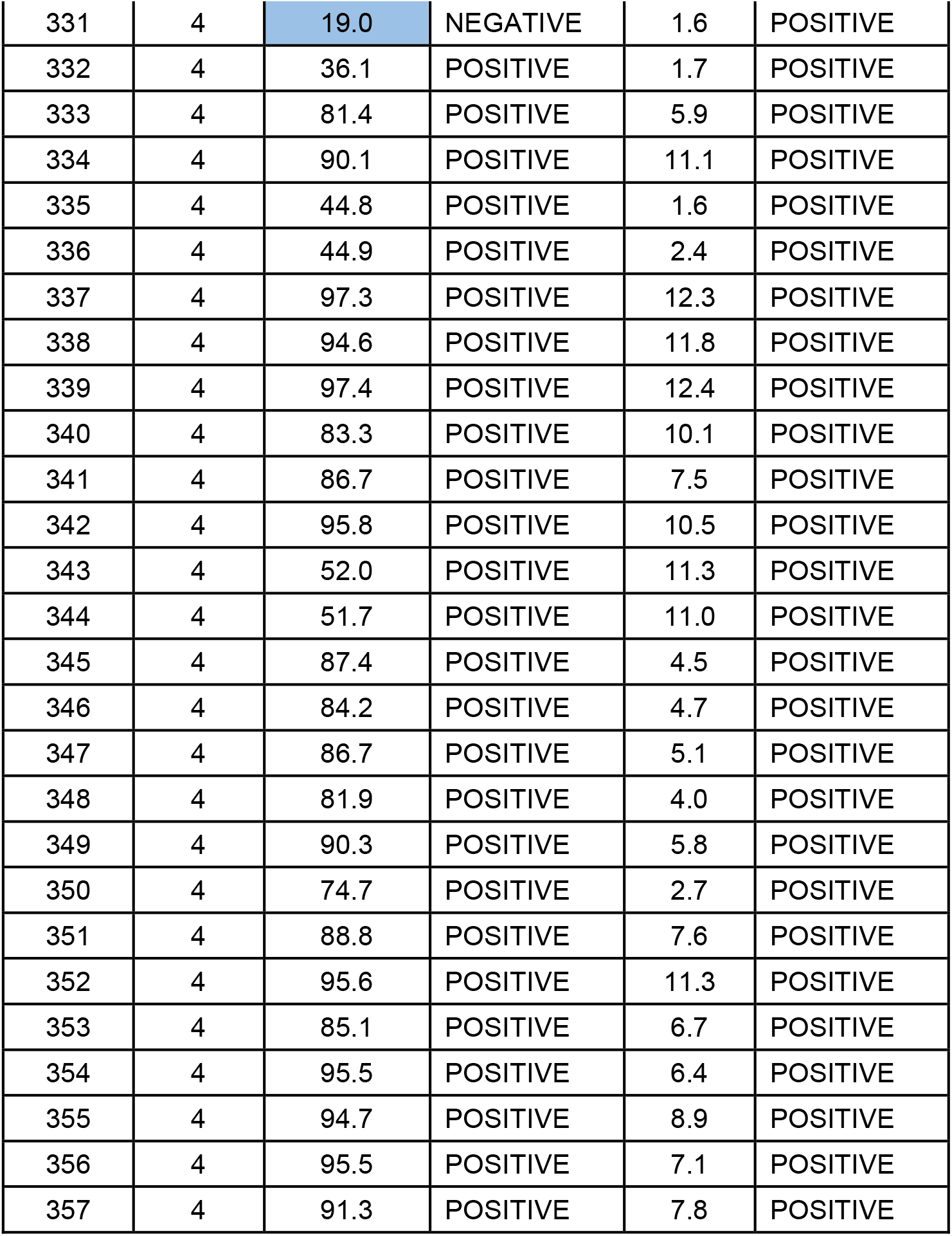
Evaluation of Neutralizing Antibodies Detection by Anti-SARS-CoV-2 DBC Serological tests. Results of testing samples from 4 sources on GenScript cPass™ SARS-CoV-2 Neutralization Antibody Detection/Surrogate Virus Neutralization Test Kit and DBC IgG anti-SARS-CoV-2 authorized serological test. Samples were sourced from (1) infected individuals PCR proven to be SARS-CoV-2 positive, (2) COVID-19 infected ICU patients, (3) fully vaccinated individuals (two doses of either Pfizer or Moderna vaccines), and (4) Reference samples from NIBSC/WHO. For the GenScript kit, a Positive outcome was considered at ≥30%. For the DBC kits, a Positive outcome was considered at a ratio of >1.2, Borderline 1.2-1.0, and Negative <1.0.

**Figure S1.**
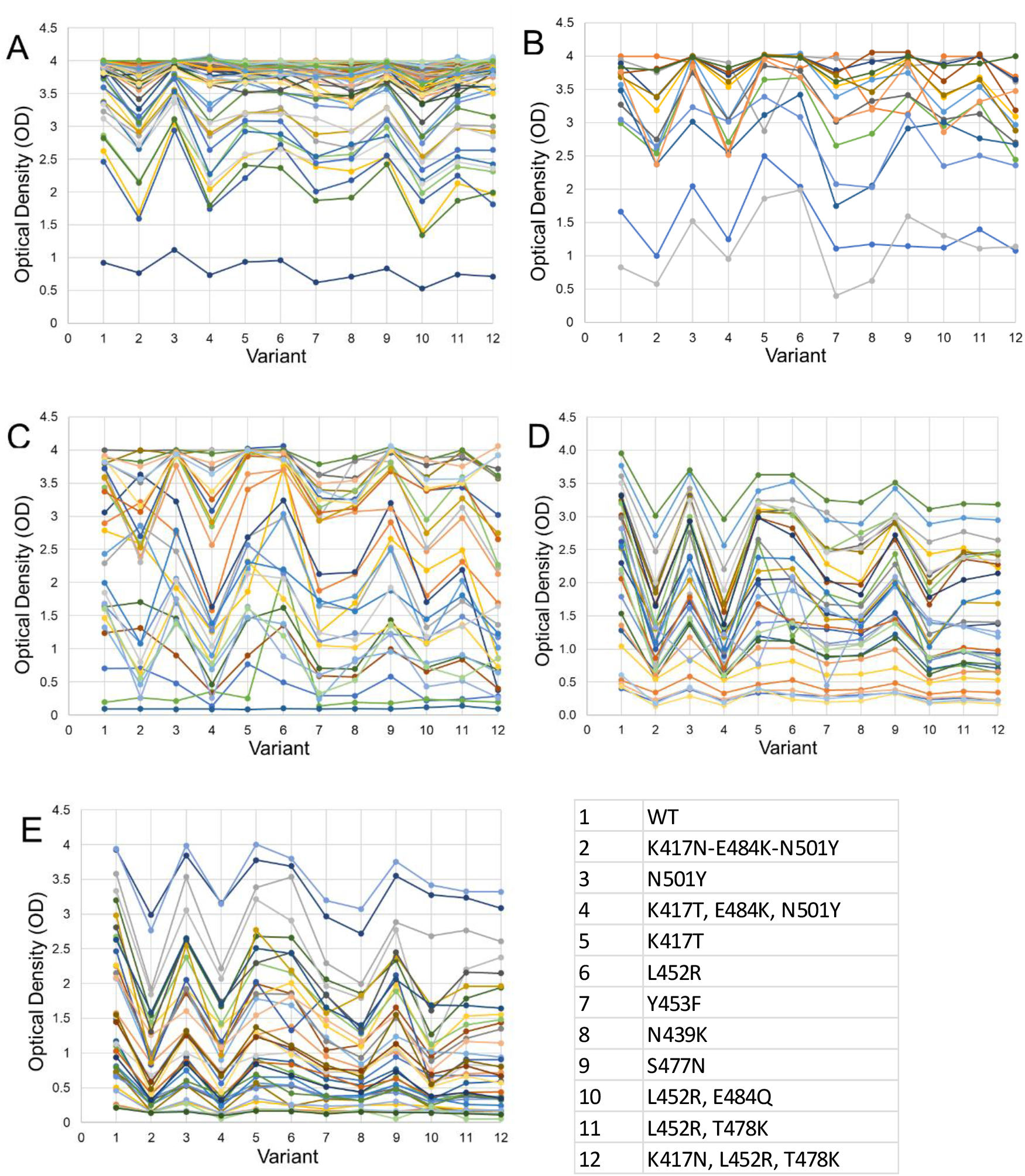
Individual variability in vaccine and convalescence sera reactivity to WT SARS-CoV-2 RBD antigen and antigens bearing mutations. Each line represents the optical density values from an individual donor. **A**. Vaccinated, **B**. ICU patients 10+ days, **C**. ICU patients 2-7 days, **D**. NIBSC/WHO (WHO) panel, **E**. Ambulatory and hospitalized (but not critically ill) population (AHP) panel.

**Figure S2.**
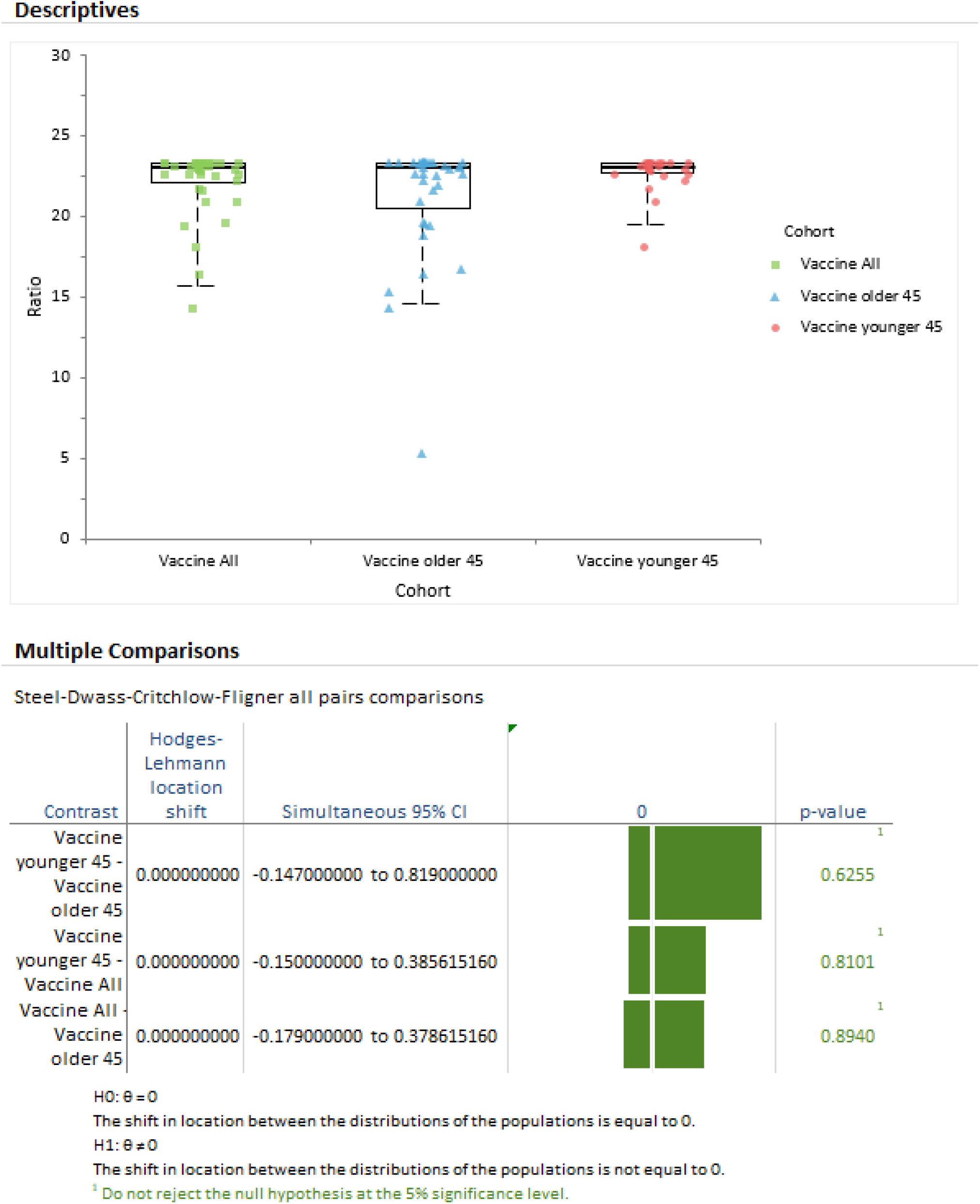
Reactivity of sera from vaccinated individuals (Vaccine) to a recombinant antigen comprising the RBD of wild type (WT) SARS-CoV-2. The vaccine results were split in those older or younger than 45 years. The median is plotted as a line. The box represents 1st to 3rd quartiles and the whiskers the minimum and maximum of the 95% confidence interval.

**Figure S3.**
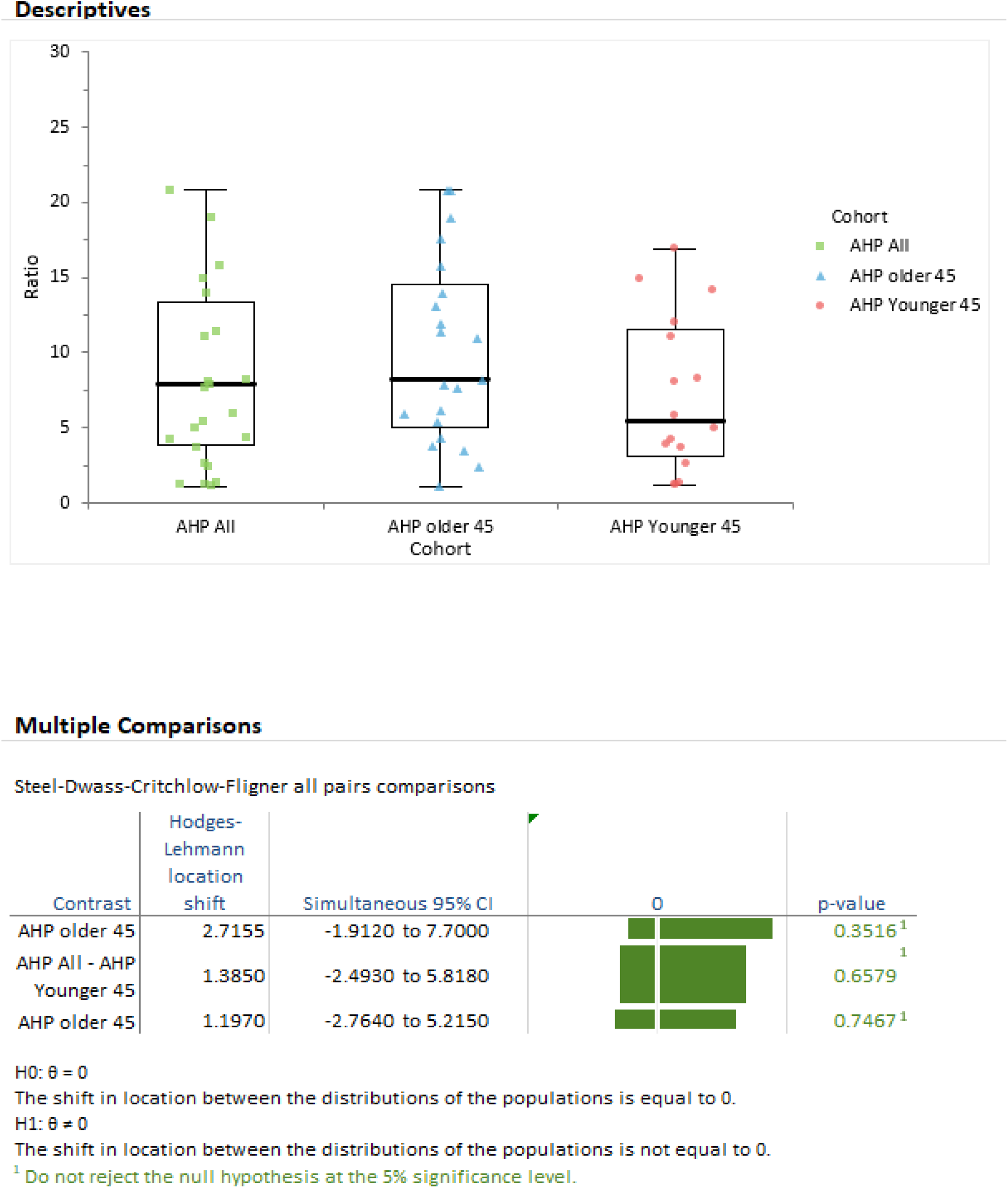
Reactivity of sera from ambulatory and hospitalized, but not critically ill, population (AHP), to a recombinant antigen comprising the RBD of wild type (WT) SARS-CoV-2. The AHP results were split in those older or younger than 45 years. The median is plotted as a line. The box represents 1st to 3rd quartiles and the whiskers the minimum and maximum of the 95% confidence interval.

## Notes

### Competing Interest Statement

All authors have completed the ICMJE uniform disclosure form at www.icmje.org/coi_disclosure.pdf and declare: PH, IH, CM, MAP, XW, JZ, JAC are currently employed by Diagnostics Biochem Canada, the manufacturer of the DBC ELISA used in this study; there are no other relationships or activities that could appear to have influenced the submitted work.

### Funding Statement

This study was partially funded by the Industrial Research Assistance Program of the National Research Council of Canada and a Grant from the National Defense of Canada.
D.D.F. received funding from Western University (Research), the Departments of Medicine and Pediatrics at Western University, the Lawson Health Research Institute (https://www.lawsonresearch.ca/), the London Health Sciences Foundation (https://lhsf.ca/), London Community Foundation and the AMOSO Innovation Fund.

### Author Declarations

Commercial samples were sourced through Access Biologicals (Vista, California, USA), Lampire Biological Laboratories (Pipersville, PA, USA), or Plasma Services Group, Inc (Moorestown, NJ, USA), each of which confirmed patient consent and participation in an Institutional Review Board (IRB) approved protocol. For critically ill patient recruitment, waived consent was approved for a short, defined period (Western University, Research Ethics Board [REB] number 1670). Samples obtained through the WHO database were originally collected under WHO protocols and ethical considerations as follows. Convalescent plasma and serum from PCR-confirmed SARS-CoV-2-infected patients was kindly donated by Coronavirus Clinical Characterisation Consortium (ISARIC4C consortium) through the University of Liverpool, UK; Papworth Hospital, Cambridge, UK; NHS Blood and Transplant, UK; and Oslo University Hospital, Norway. All patient donors gave informed consent for the use of their plasma or serum, and samples were anonymized. For material provided by ISARIC4C, ethical approval was given by the South Central-Oxford C Research Ethics Committee in England (reference 13/SC/0149), and by the Scotland A Research Ethics Committee (reference 20/SS/0028). The study was registered at https://www.isrctn.com/ISRCTN66726260. The NIBSC Human Material Advisory Committee (project 16/005MP) approved this project. The authors confirm that all necessary patient/participant consent has been obtained and the appropriate institutional forms have been archived, and that any patient/participant/sample identifiers included were not known to anyone outside the research group or have been sufficiently anonymized and so cannot be used to identify individuals.

